# Pregnancy and the prognosis of patients previously treated for differentiated thyroid cancer: a systematic review and meta-analysis

**DOI:** 10.1101/2023.03.11.23287150

**Authors:** Rui Shan, Xin Li, Wu-Cai Xiao, Jing Chen, Fang Mei, Shi-Bing Song, Bang-Kai Sun, Zheng Liu

**Affiliations:** Department of Maternal and Child Health, School of Public Health, Peking University; Department of General Surgery, Peking University Third Hospital; Department of Pathology, Peking University Third Hospital, School of Basic Medical Sciences, Peking University Health Science Center; Information Management and Big Data Center, Peking University Third Hospital

## Abstract

**IMPORTANCE:** Differentiated thyroid cancer (DTC) is commonly diagnosed in women of child-bearing age, but whether pregnancy influences the prognosis of DTC remained controversial.

**OBJECTIVE:** This systematic review and meta-analysis aimed to summarize and appraise the existing evidence of the impact of pregnancy on the prognosis of patients previously treated for DTC.

**DATA SOURCES:** We searched PubMed, Embase, Web of Science, Cochrane, and Scopus until February 2023.

**STUDY SELECTION:** Studies of patients diagnosed and treated for DTC before pregnancy reporting the recurrence/progression condition of DTC were included. Case reports and studies failing to identify the time of diagnosis or initial treatment were excluded.

**DATA EXTRACTION AND SYNTHESIS:** Meta-analyses were conducted according to MOOSE guideline. Data extraction was conducted by two independent investigators with a standard form. Pooled effect estimates were calculated in a random-effects model.

**MAIN OUTCOMES AND MEASURES:** DTC recurrence/progression and the type of recurrence/progression (structural or biochemical).

**RESULTS:** Among the 10 included studies (n = 625), 4 (n = 143) of them compared the pregnancy group with the non-pregnancy group while the remaining 6 (n = 482) only included the pregnant patients. The pooled proportion of recurrence/progression in all pregnant patients was 13% (95% CI, 6%, 25%). Compared with the non-pregnancy group, the pooled odds ratio of recurrence/progression in the pregnancy group was 0.75 (95% CI, 0.45, 1.23). Two included studies focused on patients with distant metastasis and also did not observe difference in disease recurrence/progression between the pregnancy group and the non-pregnancy group [OR, 0.51 (95% CI, 0.14-1.87)]. Six included studies also reported response to therapy status prior to pregnancy, and the pooled proportion for recurrence/progression in pregnant DTC patients with excellent response (n=287), indeterminate response (n=44), biochemical incomplete response (n=41) and structural incomplete response (n=70) was 0.00 (95% CI, 0.00-0.86), 0.09 (95% CI, 0.00-0.99), 0.20 (95% CI, 0.06-0.46) and 0.45 (95% CI, 0.17-0.76), respectively. There was a trend for an increasingly higher risk of recurrence/progression from excellent, indeterminate, biochemical incomplete to structural incomplete response to therapy (*P*<0.05).

**CONCLUSIONS AND RELEVANCE:** Pregnancy appears to have a minimal impact on the prognosis of DTC with initial treatment. Clinicians may pay more attention to the progression of DTC among pregnant women with biochemical and/or structural persistence.

**Key Points:** *Question:* Does subsequent pregnancy has an impact on the prognosis of patients previously treated for differentiated thyroid cancer (DTC)?

*Findings:* In this systematic review and meta-analysis of 10 studies including 625 patients previously treated for DTC and underwent pregnancy subsequently, pregnancy might have a minimal impact on DTC recurrence/progression. Patients with biochemical and/or structural incomplete response to DTC treatment prior to pregnancy appears to have a higher risk of DTC recurrence/progression compared to those with excellent or indeterminate response.

*Meaning:* Though pregnancy appears to have little influence on the prognosis of patients previously treated for DTC, patients with biochemical and/or structural persistence should be more carefully monitored during pregnancy.

## Introduction

Up to 2020, thyroid cancer has become the fifth most common cancer in women^1^. The incidence rate of thyroid cancer was almost 2.6 times higher in females compared to that in males^2^. According to SEER Statistics Review, the female age-adjusted incidence rates of thyroid cancer have risen from 6.15/10^5^ in 1980 to 19.72/10^5^ in 2017^3^. From 2005 to 2015, the female age-standardized incidence rates of thyroid cancer have increased from 6.68/10^5^ to 20.28/10^5^ in China^4^. About 95% of thyroid cancer consists of the type of differentiated thyroid cancer (DTC)^5^.

DTC is very common in women of childbearing age, among whom almost 14.4/10^5^ females were diagnosed around pregnancy^6^. There are hypotheses suggesting the increasing release of hormone of estrogen and human chorionic gonadotropin during pregnancy might stimulate the recurrence/progression of thyroid cancer. Human chorionic gonadotropin has a similar function like thyroid-stimulating hormone or thyrotropin, provoking thyroid hormones secretion which contribute to the rise of estrogen levels^7^. Via MAPK cytoplasmic signaling, estrogen may affect proliferation and growth of thyroid cancer cells^8^, leading to potentially poorer prognosis. Thus, whether pregnancy was indeed related to disease progression has drawn great attention of both clinicians and patients with DTC.

Relevant studies were heterogeneous in study design, population, outcome measures, and study findings. But to our knowledge, only one systematic review has focused on the prognosis of DTC associated with pregnancy. This systematic review only included 4 studies designed with a comparison group and the included patients were diagnosed during pregnancy from 1994 to 2010.

One of included studies suggested a higher rate of recurrence in patients with DTC related to pregnancy; however, the rest of three studies showed no difference in prognosis of pregnancy-related DTC as compared to control groups. Subsequently, more studies have been conducted after this systematic review. Up to now, two Italian studies reported significantly higher risk of recurrence in patients who were diagnosed with DTC during pregnancy or soon after delivery^8,9^. As for DTC patients diagnosed and treated before pregnancy, a Chinese study claimed pregnancy has no impact on prognosis^10^. Discrepancy across studies may be attributed to the varied time of diagnosis (during pregnancy or prior to pregnancy), study design (single-group study or study with comparison group), or patients with different response to therapy status. Moreover, there has been no systematic review quantitatively synthesizing the association of pregnancy with prognosis of previously-treated DTC patients by using the meta-analytic technique.

To fill the research gap, this study aimed to conduct a systematic review and meta-analysis to quantitatively summarize the most up-to-date studies and answer whether pregnancy was associated with the prognosis of patients previously treated for DTC. We also performed several subgroup and sensitive analyses to elucidate the potential influence of study design and response to therapy status on the main study findings.

## Methods

### Search Strategy

The present study was conducted according to MOOSE guidelines and was registered on PROSPERO (CRD42022367896). Our research systematically searched citation databases including PubMed, Embase, Web of Science, Cochrane and Scopus, combining MESH terms of “Thyroid Cancer” and “Pregnancy” (Appendix A). All studies before February 1^st^ 2023 were included.

### Inclusion and Exclusion Criteria for Studies

The inclusion criteria for citations was based on the PICOS elements: (1) population: patients diagnosed with DTC before pregnancy; (2) intervention: patients initially treated (operation with/without RAI treatment) before pregnancy; (3) comparators: studies designed with or without a comparison group; (4) outcomes: studies reported recurrence/progression of DTC; (5) study design: case-control design or case series design. The exclusion criteria for citations: (1) case reports; (2) studies with unclear information (fail to identify the time of diagnosis or initial treatment); (3) published in languages other than English; (4) abstracts and unpublished studies.

### Quality assessment

The predefined U.S. Preventive Services Task Force criteria was applied to assess the quality of studies^11^. Studies were rated as “good”, “fair”, or “poor” according to the study design (Appendix B and Appendix C). The quality assessment tables were designed and revised into 3 aspects for studies with a comparison group and 2 for single-group studies based on Chou R 2022, respectively. We considered age at pregnancy and TNM stage/ATA risk stratification/response-to-therapy status as the primary confounders and influencing factors. For studies with a comparison group, (1) groups should be comparable in influencing factors such as age at pregnancy and TNM stage/ATA risk stratification/response-to-therapy status at baseline; (2) confounders should be accurately ascertained and adequately controlled by multifactor analyses; (3) studies should prespecify and define outcomes, and use accurate methods. Studies that met the all 3 aspects were rated as “good”, those met 2 aspects were graded as “fair”, and those only met 1 aspect or less were rated as “poor”. As for single-group studies, those (1) accurately ascertained confounders, (2) predefined outcomes and used accurate methods were rated as “fair”, while those only met 1 aspect or less were graded as “poor”.

### Data extraction

Two researchers (RS; XL) with rich experiences in public health and general surgery extracted the following information from the included studies: (1) basic characteristics: study design, country, enrollment, age, interval time between diagnosis and pregnancy, interval time between initial treatment and pregnancy, interval time between initial treatment and pregnancy, follow-up time, histology, conditions of extrathyroidal extension, lymph node metastasis and distant metastasis at baseline; (2) primary outcomes: number (proportion) of recurrence/progression, type of recurrence/progression (structural or biochemical).

### Definitions of recurrence, risk stratification and response to therapy status

Structural recurrence/progression was defined as appearance of new local/metastasis lesions, or 20% or more increase in size of a pre-pregnancy lesion^12,13^. Biochemical recurrence/progression was defined as 20% or more increase in serum thyroglobulin (Tg) from the pre-pregnancy level and/or consistent increase of 20% or more in serum anti-Tg antibodies^10,12^.

Response to therapy status was classified into four categories: excellent response, indeterminate response, biochemical incomplete response, and structural incomplete response, defined by 2015 ATA^14^ and Momesso et al (2016)^15^.

### Data analysis

Data analysis was conducted in R (version 4.2.1). Meta-analyses of single-proportions for recurrence/progression in each study was calculated in generalized linear mixed model. We pooled odds ratio (OR) to compare the proportion of disease recurrence/progression in the pregnancy group with that in the non-pregnancy group by using Mantel-Haenszel method with random effects variants (“metabin” function in R package “meta”). Additionally, two subgroup analyses were conducted based on type of recurrence (structural or biochemical) and response to therapy status by using generalized linear mixed model. We used Chi-square test to examine whether the association of pregnancy with disease progression differed by the subgroup variable. For subgroup analysis based on 4 levels of response to therapy status, we also conducted a Trend test by using meta-regression.

## Results

### Study selection

A total of 5366 studies were identified from databases (PubMed: 720, Cochrane: 3, Embase: 869, Web of Science: 1747, Scopus: 1927). 2098 duplicate studies were removed. After screening, 38 studies were assessed for eligibility, from which 28 studies that failed to meet inclusion criteria were excluded. Ultimately, 10 studies were included, consisting of 4 studies with a comparison group^10,13,16,17^ and 6 single-group studies^12,18-22^ (Figure 1).

**Figure 1.**
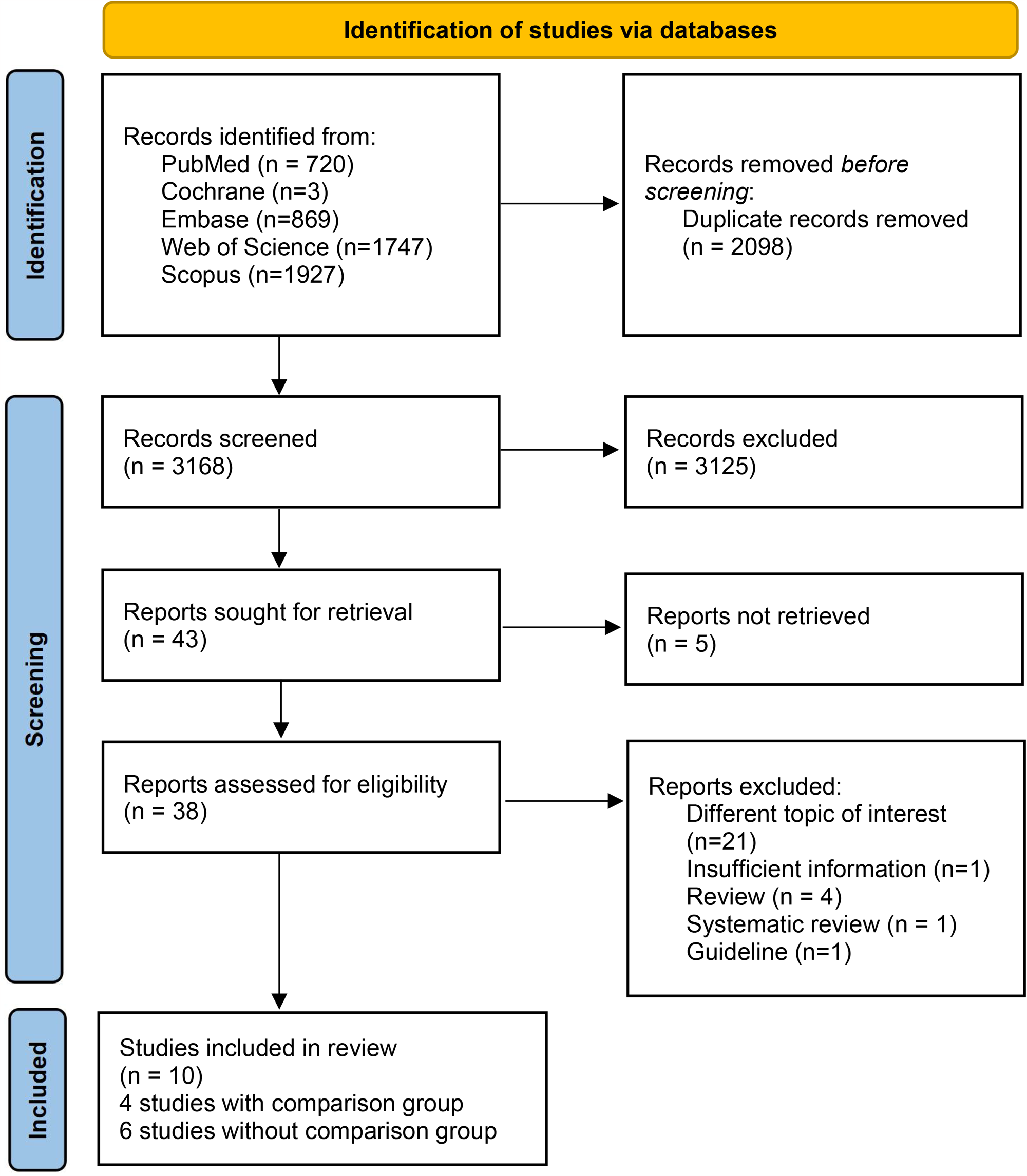
Literature flow diagram.

### Study characteristics

The study characteristics of ten studies was shown in Table 1 and the pathological characteristics in Table 2. Among the ten included studies, four of them were designed with a comparison group while the remaining six were single-group designed. Sample size of the ten studies ranged from 8 to 235 with age at pregnancy or delivery basically 28-35 years. The follow-up time varied from 0.1 to 36 years.

**Table 1.** Study Characteristics.

| Study | Study design | Country | Enrollment, n | Age at pregnancy or delivery, Mean ± SD, years | Interval time between diagnosis and pregnancy, Mean ± SD | Interval time between initial treatment and pregnancy, Mean ± SD | Follow-up, Mean ± SD |
| --- | --- | --- | --- | --- | --- | --- | --- |
| Leboeuf R et al, 2007<sup>19</sup> | Single-group study | USA | 36 | 33.6 ± 3.9<br>(Median, 33.5; Range, 24.3-42.2) | NR [a] | 5.5 ± 3.8 years<br>(Median, 4.3) | Median 4 months (Range 0.1-1.7 years) |
| Rosario PW et al, 2007<sup>22</sup> | Single-group study | Brazil | 64 | 32 (Range, 23-39) | NR | 30 months<br>(Range, 15-74) | NR |
| Hirsch D et al, 2010<sup>12</sup> | Single-group study | Israel | 63 | 29.7 ± 4.7<br>(Median, 29.41; Range, 20.1-41.2) | NR | 5.08 ± 4.39 years (Median, 3.20) [b] | 4.84 ± 3.8 years<br>(Range, 3 months to 17.3 years;<br>Median, 3.75 years) |
| Budak A et al, 2013<sup>16</sup> | Study with a comparison group | Turkey | **Pregnant**, n=36<br>**Control**, n=36 | **Pregnant**,<br>29.9 ± 5.1<br>(Range, 21-40) | 40.5 ± 26.4 months<br>(Range, 12-120) | NR | **Pregnant**, 6.05 ± 2.5 years<br>**Control**, 5.2 ± 0.5 years |
| Rakhlin L et al, 2017<sup>21</sup> | Single-group study | USA | 235 | 34 ± 0.4<br>(Median, 34; Range, 20-45) | Mean, 4.9 ± 0.3 years<br>(Median, 3; Range, 0.1-23) | NR | 3-12 months |
| Driouich Y et al, 2021<sup>17</sup> | Study with a comparison group | Morocco | **Pregnant**, n=42<br>**Control**, n=75 | **Pregnant**,<br>35.0 ± 6.5 [c] | NR | 4.4 ± 3.1 years | **Pregnant**, 14.45 ± 2.4 months<br>**Control**, 18.24 ± 3.7 months |
| Nobre GM et al, 2021<sup>20</sup> | Single-group study | Brazil | 76 | 32 (Range, 21-44) | Median, 4.8 years (Range, 0.9-19) [d] | NR | 11 years (Range, 1-47) |
| Xi C et al, 2021<sup>10</sup> | Study with a comparison group | China | **Pregnant**, n=37<br>**Control**, n=87 | **Pregnant**,<br>28.59 ± 4.086<br>(Median, 28; Range, 22-40) | NR | 46.27 ± 25.74 months<br>(Median, 42; Range, 7-102) | Median, 82 months<br>(Range, 25-136) |
| Colombo C et al, 2022<sup>18</sup> | Single-group study | Italy | 8 | 33.1 ± 4.6<br>(Median, 35; Range, 27-38) | Mean, 62 ± 43.8 months<br>(Median, 66; Range, 12-120) | NR | Mean, 97 months |
| Yamazaki H et al, 2022<sup>13</sup> | Study with a comparison group | Japan | **Pregnant**, n=28<br>**Control**, n=97 | **Pregnant**,<br>32 (Range, 25-45) | Median, 4.8 years (Range, 0.3–18.9) | Median, 2.1 years (Range, 0.5–14.5) | **Pregnant**, Median 10.9 years<br>(Range, 4.2–32.5)<br>**Control**, Median 6.6 years<br>(Range, 0.1–36.0) |

[a]: NR, Not Reported
[b]: Interval time between initial treatment and delivery
[c]: Age at diagnosis
[d]: Interval time between diagnosis and delivery

**Table 2.** Pathological Characteristics.

| Study | Study design | Histology | Extrathyroidal extension | Lymph node metastasis | Distant metastasis |
| --- | --- | --- | --- | --- | --- |
| Leboeuf R et al, 2007<sup>19</sup> | Single-group study | Papillary 66.7%<br>Follicular variant of PTC 27.8%<br>Follicular 2.8%<br>Poorly differentiated 2.8% | NR* | 17 (56.7%) | 1 (3.1%) |
| Rosario PW et al, 2007<sup>22</sup> | Single-group study | PTC 75%<br>FTC 25% | NR | NR | NR |
| Hirsch D et al, 2010<sup>12</sup> | Single-group study | PTC | 1 (1.6%) | 31 (49.2%) | 2 (3.2%) |
| Budak A et al, 2013<sup>16</sup> | Study with a comparison group | **Pregnant and Control,**<br>PTC 94.4%; FTC, 5.6% | NR | **Pregnant, 2 (6%)**<br>**Control, 2 (6%)** | **Pregnant, 3 (8%)**<br>**Control, 1 (3%)** |
| Rakhlin L et al, 2017<sup>21</sup> | Single-group study | Classic papillary 73.6%<br>Follicular variant, papillary 19.6%<br>Poorly differentiated 3.4%<br>Follicular 2.6%<br>Hürthle cell 0.9% | NR | 115 (48.9%) | 10 (4.3%) |
| Driouich Y et al, 2021<sup>17</sup> | Study with a comparison group | PTC | **Pregnant, 4(10%)**<br>**Control, 5(7%)** | **Pregnant, 10(24%)**<br>**Control, 35(46%)** | **Pregnant, 3(25%)**<br>**Control, 18(24%)** |
| Nobre GM et al, 2021<sup>20</sup> | Single-group study | PTC 76%<br>FVPTC 18%<br>FTC 2.6%<br>Other 2.6% | NR | 40 (55%) | 7 (9.2%) |
| Xi C et al, 2021<sup>10</sup> | Study with a comparison group | **Pregnant,**<br>PTC 86.47%; FTC 3.51%<br>**Control,**<br>PTC 88.51%; FTC 11.49% | **Pregnant, 9 (24.32%)**<br>**Control, 34 (37.93%)** | **Pregnant, 32 (86.5%)**<br>**Control, 78 (89.7%)** | **Pregnant, 34 (91.9%, lung); 3 (8.11%, lung and bone)**<br>**Control, 84 (99.6%, lung); 3 (3.4%, lung and bone)** |
| Colombo C et al, 2022<sup>18</sup> | Single-group study | Conventional variant PTC 87.5%<br>Sclerosing variant PTC 12.5% | NR | 5(62.5%, neck) | 2 (25%, lung) |
| Yamazaki H et al, 2022<sup>13</sup> | Study with a comparison group | **Pregnant,**<br>PTC 82%; FTC 18%<br>**Control,**<br>PTC 88%; FTC 12% | NR | **Pregnant, 20%**<br>**Control, 65%** | **Pregnant,**<br>26 (93%, lung); 2 (7%, lung+bone)<br>**Control,**<br>86 (88%, lung); 7 (8%, bone); 3 (3%, lung+bone); 1 (1%, other) |

\*: NR, Not Reported

### Pregnancy and prognosis of DTC

All of ten studies were included in meta-analysis. In total, ten studies reported 625 patients who underwent pregnancy after diagnosis and initial treatment for DTC, 101 of whom experienced recurrence. The proportion of recurrence/progression in random effects model was 13% (95% CI, 6%-25%; I^2^=58%) in pregnant patients previously treated for DTC (Supplementary Figure 1).

Specifically, there were four studies classifying recurrence into structural and biochemical types. The proportion of structural and biochemical recurrence/progression in pregnant DTC patients was 0.06 (95% CI, 0.03-0.11; I^2^=0%) and 0.10 (95% CI, 0.04-0.23; I^2^=62%), respectively (Supplementary Figure 2). We observed no evidence of difference between the proportion of structural recurrence/progression and the proportion of biochemical recurrence/progression (*P* = 0.09).

**Figure 2.**
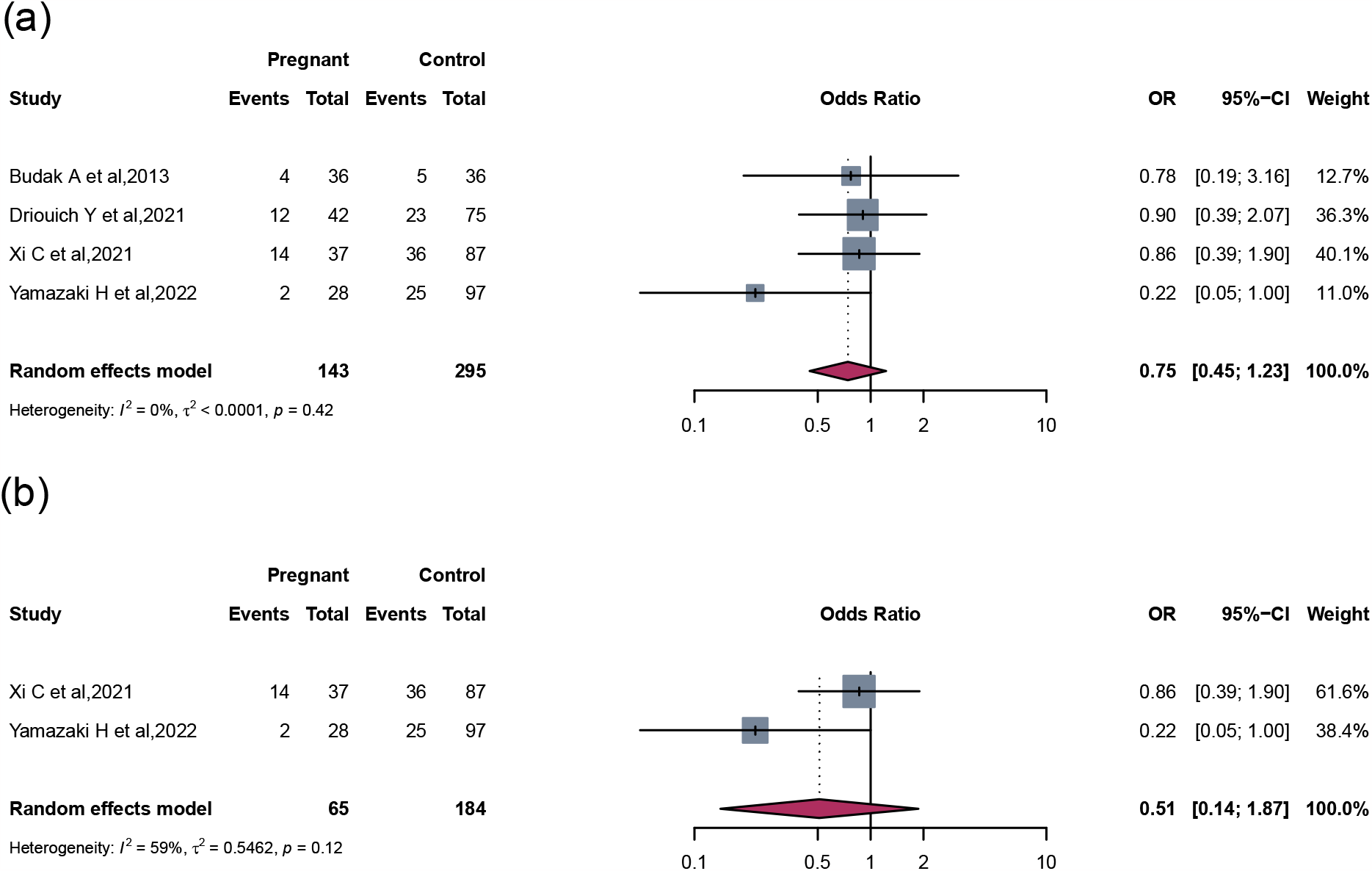
The pooled recurrence/progression (OR) (a) between pregnant patients and non-pregnant controls previously treated for DTC and (b) in patients with distant metastasis at the time of diagnosis.

Among 4 of the 10 included studies designed with a comparison group, we further explored whether the risk of recurrence/progression differed between the pregnant group and non-pregnancy group. The OR for recurrence/progression in pregnant DTC patients was 0.75 (95% CI, 0.45-1.23; I^2^=0%) as compared to non-pregnant patients (Figure 2a).

Specifically, the included two studies focused on DTC patients with distant metastasis at diagnosis. The OR for recurrence/progression in DTC patients with distant metastasis who became pregnant later was 0.51 (95% CI, 0.14-1.87; I^2^=59%) compared with those who didn’t get pregnant (Figure 2b).

### Response to therapy status and prognosis of DTC

There were six studies described the response to therapy status before pregnancy. The pooled proportion for recurrence/progression was 0.00 (95% CI, 0.00-0.86) in pregnant DTC patients with excellent response (n=287), 0.09 (95% CI, 0.00-0.99) in those with indeterminate response (n=44), 0.20 (95% CI, 0.06-0.46) in those with biochemical incomplete response (n=41), and 0.45 (95% CI, 0.17-0.76) in those with structural incomplete response (n=70) (Figure 3). Differences between four subgroups were significant (*P* = 0.01). There was a trend for a higher risk of recurrence/progression from excellent response, indeterminate response, biochemical incomplete response to structural incomplete response (Trend test, *P* < 0.05).

**Figure 3.**
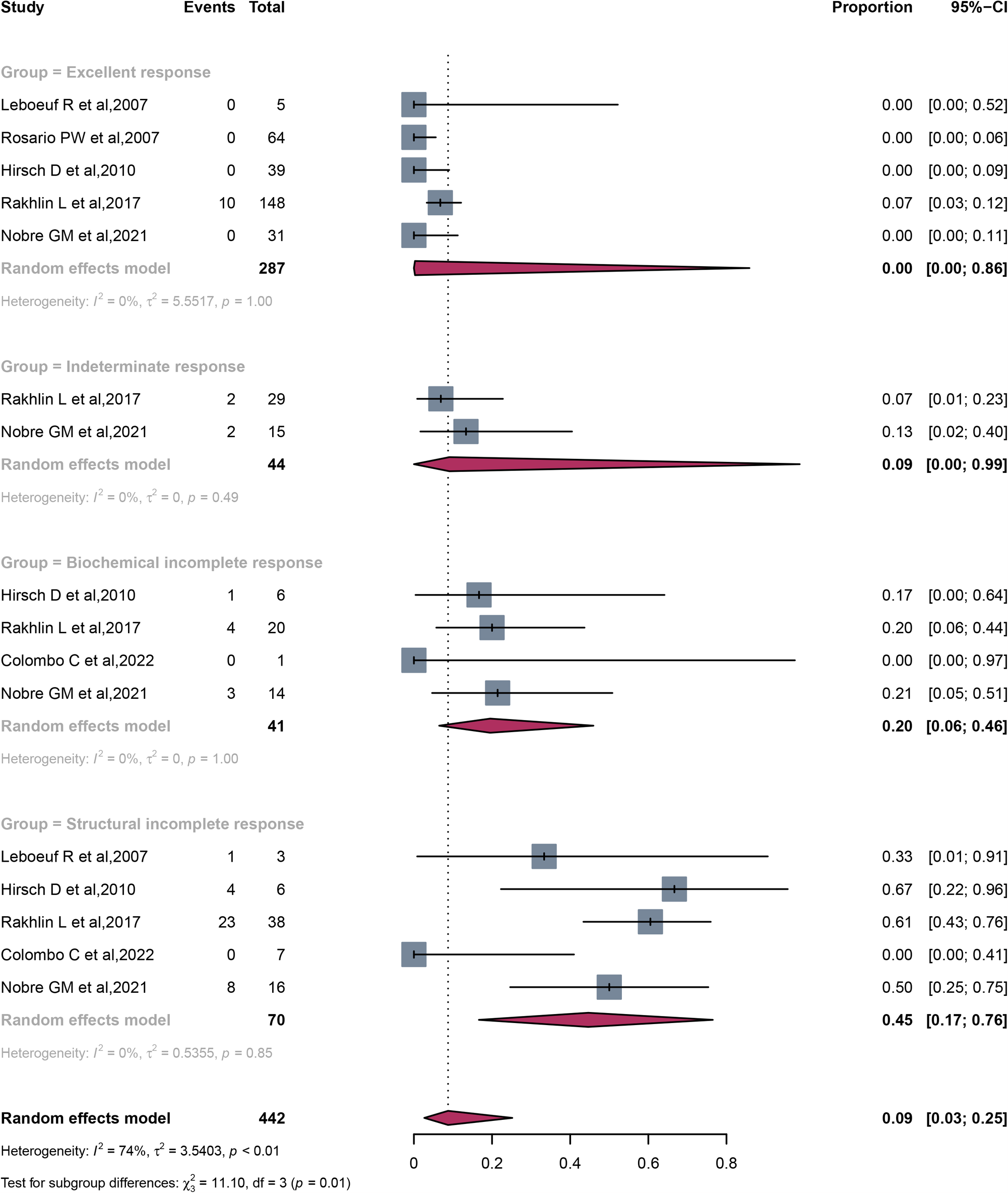
Response to therapy status and the pooled recurrence/progression in pregnant patients.

## Discussion

Our study was among the first to systematically review and quantitatively summarize the potential influence of pregnancy after the treatment of DTC on disease progression. We conducted several sensitivity and stratification analyses and the results consistently indicated that pregnancy might have a minimal impact on prognosis of DTC with initial treatment before pregnancy.

Meta-analysis results showed that, the pooled proportion of recurrence/progression in all the pregnant women of included studies is relatively small [13% (95% CI, 6%-25%)], since the recurrence/progression rate in the non-pregnant control groups was around 14-41%. The fact indicated that pregnancy only has a minor impact on stimulating recurrence/progression. For the 4 included studies with a comparison group, the risk of recurrence/progression of DTC in the pregnancy women showed no evidence of different from those who didn’t underwent pregnancy. Even in DTC patients with distant metastasis, the pooled results of the included 2 studies still revealed no evidence of pregnancy increasing the risk of recurrence/progression.

To our knowledge, there was only one systematic review conducted in 2011^23^. But this study included only 4 studies and the patients were diagnosed during pregnancy or within 12 months postpartum. None of previous reviews conducted quantitative analyses, making it difficult to reach a conclusion about the association of pregnancy with DTC progression. The research question becomes even complicated considering potential confounders of the varied time of diagnosis (during pregnancy or prior to pregnancy), study design (single-group study or study with comparison group), and different response to therapy status. As such, our study specifically included previously-treated DTC patients prior to pregnancy, conducted sensitivity analyses among patients with distant metastasis, and subgroup analysis were performed based on response to therapy status.

Our finding of little association between pregnancy with DTC progression should be interpretated cautiously. None of the included studies explicitly compared health condition between the pregnant group and the non-pregnant group. Thus, we could not exclude the possibility that pregnant women were in relatively good health condition and this might dilute the association of pregnancy with DTC progression.

### Limitations and strengths

Our systematic review and meta-analysis had limitations. First, we only searched citations published in English. Second, we didn’t assess potential publication bias since small number of studies. Third, almost all included studies were retrospective, leading to potential confounding, selection, information, temporal and immortal time bias^24^. Nevertheless, our study fully searched database of PubMed, Embase, Web of Science, Cochrane and Scopus and synthesized outcomes by meta-analyses quantitively. Our study is the first to systematically explore the impact of pregnancy on previously-treated DTC patients. Furthermore, we performed several subgroup and sensitive analyses to excavate the effect of response to therapy status prior to pregnancy and original disease stage (with or without distant metastasis) on disease recurrence/progression, which verified the robustness of conclusion and made it more persuasive.

### Research and clinical implications

Based on findings of our review, we suggested future studies to increase the sample size due to the insufficient statistical power of the existing population scale (Range, n=8-235). Additionally, we suggested future studies to design the study by comparing the pregnancy group with the non-pregnancy group. In this way, we could better elucidate whether the DTC recurrence/progression was due to pregnancy process itself or simply a natural process of DTC in both pregnant and non-pregnant patients. To form a study of high quality, confounding factors like age at pregnancy and TNM stage/ATA risk stratification/ response-to-therapy status were required to be accurately ascertained and adequately controlled. Multivariate analyses were needed to adjust other factors and demonstrate the independent risk factors. Besides, the definition of recurrence/progression should be ascertained and prespecified. It was favorable if the type of recurrence/progression (structural or biochemical) was also clearly distinguished and defined.

Our study also had clinical implications. Clinicians should provide personalized suggestions to patients based on their response to therapy status before pregnancy. For patients with incomplete response to therapy prior to pregnancy, active monitoring of disease progression is strongly suggested during pregnancy; for those with excellent or indeterminate response, pregnancy might not need to be postponed and actively monitored^25^.

## Conclusions

Our up-to-date systematic review showed that pregnancy appears to have a minimal impact on the prognosis of DTC with initial treatment. Clinicians may pay more attention to progression of DTC among pregnant women with biochemical and/or structural persistence.

## Supporting information

Appendix A search strategy

Appendix B with a comparison group

Appendix C single group

## Data Availability

All data produced in the present work are contained in the manuscript

