## Supplementary figures and images for "Pregnancy and the prognosis of patients previously treated for differentiated thyroid cancer: a systematic review and meta-analysis"

### Supplementary Figure 1

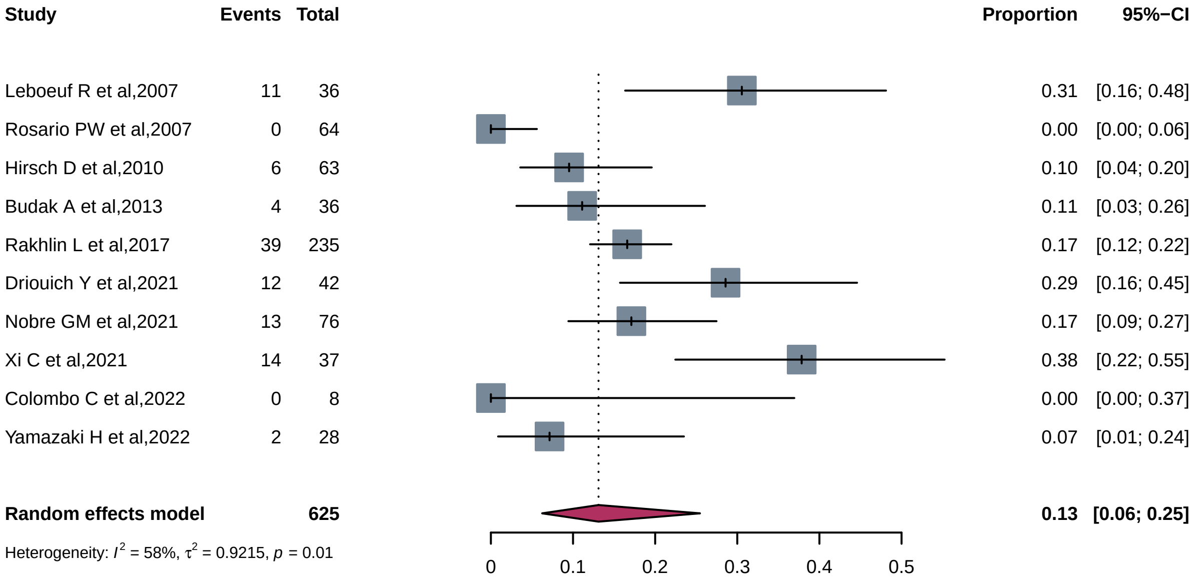
