## Supplementary Figure 2 for "Pregnancy and the prognosis of patients previously treated for differentiated thyroid cancer: a systematic review and meta-analysis"

| Study | Events | Total | Proportion | 95%−CI |
|---|---|---|---|---|
| Group = Structural recurrence | | | | |
| Leboeuf R et al,2007 | 3 | 36 | 0.08 | [0.02; 0.22] |
| Hirsch D et al,2010 | 4 | 63 | 0.06 | [0.02; 0.15] |
| Rakhlin L et al,2017 | 11 | 235 | 0.05 | [0.02; 0.08] |
| Nobre GM et al,2021 | 6 | 76 | 0.08 | [0.03; 0.16] |
| **Random effects model** | | **410** | **0.06** | **[0.03; 0.11]** |
| Heterogeneity: $I^2 = 0\%$, $\tau^2 = 0$, $p = 0.67$ | | | | |
| Group = Biochemical recurrence | | | | |
| Leboeuf R et al,2007 | 8 | 36 | 0.22 | [0.10; 0.39] |
| Hirsch D et al,2010 | 2 | 63 | 0.03 | [0.00; 0.11] |
| Rakhlin L et al,2017 | 28 | 235 | 0.12 | [0.08; 0.17] |
| Nobre GM et al,2021 | 7 | 76 | 0.09 | [0.04; 0.18] |
| **Random effects model** | | **410** | **0.10** | **[0.04; 0.23]** |
| Heterogeneity: $I^2 = 62\%$, $\tau^2 = 0.1943$, $p = 0.05$ | | | | |
| **Random effects model** | | **820** | **0.08** | **[0.05; 0.13]** |
| Heterogeneity: $I^2 = 60\%$, $\tau^2 = 0.1727$, $p = 0.01$ | | | | |
| Test for subgroup differences: $\chi^2_1 = 2.93$, df = 1 ($p = 0.09$) | | | | |

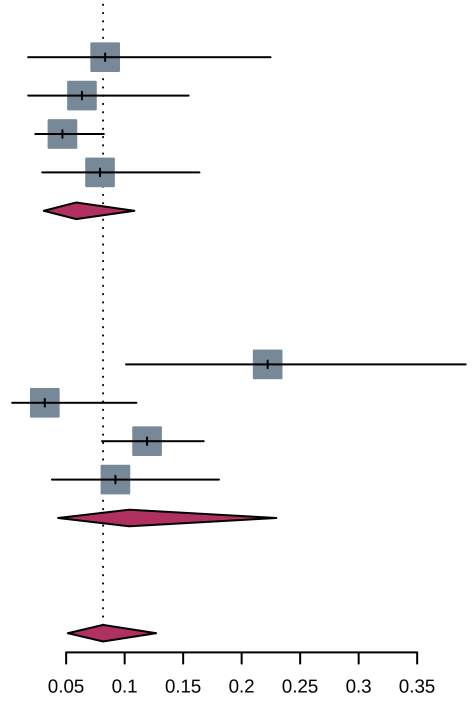
