## Appendix A search strategy for "Pregnancy and the prognosis of patients previously treated for differentiated thyroid cancer: a systematic review and meta-analysis"

**PubMed**

("Thyroid Neoplasms"[Mesh] OR "Thyroid Cancer, Papillary"[Mesh] OR "Thyroid cancer, Hurthle cell" [Supplementary Concept] OR "Thyroid Carcinoma, Anaplastic"[Mesh] OR "Thyroid cancer, medullary" [Supplementary Concept] OR "Familial medullary thyroid carcinoma" [Supplementary Concept] OR "Thyroid cancer, follicular" [Supplementary Concept]) AND "Pregnancy"[Mesh]

720 results

**Cochrane**

ID	Search	Hits

#1	MeSH descriptor: [Thyroid Neoplasms] explode all trees	833

#2	MeSH descriptor: [Pregnancy] explode all trees	28536

#3	#1 AND #2	3

**Embase**

Session Results

.......................................................

No. Query Results	Results Date

#3. #1 AND #2	869 1 Feb 2023

#2. 'pregnancy'/exp OR 'conception':ab,kw,ti	891,528 1 Feb 2023

#1. 'thyroid cancer'/exp OR 'thyroid cancer' OR	89,692 1 Feb 2023

'thyroid carcinoma':ab,kw,ti

**Web of Science**

((TS=(thyroid cancer)) OR TS=(thyroid carcinoma)) AND (TS=(pregnancy) OR TS=(conception))

1747 results

**Scopus**

(TITLE-ABS-KEY (thyroid AND cancer ) OR TITLE-ABS-KEY ( thyroid AND carcinoma ) AND TITLE-ABS-KEY ( pregnancy ) )

1927 results
