## Appendix B with a comparison group for "Pregnancy and the prognosis of patients previously treated for differentiated thyroid cancer: a systematic review and meta-analysis"

**Appendix B. Quality Assessment –Studies With a Comparison Group, Pregnancy Vs. Non-pregnancy**

| **Study** | **Groups** **comparable in age at pregnancy and** **TNM stage/ATA risk stratification/ response-to-therapy status at baseline?** | **Outcomes pre-specified and defined, and ascertained using accurate methods?** | **Confounders accurately ascertained and adequately controlled?** | **Quality Rating** |
| --- | --- | --- | --- | --- |
| Budak A et al, 2013 | Yes | No | No | Poor |
| Driouich Y et al, 2021 | Yes | Yes | Yes | Good |
| Xi C et al, 2021 | Yes | Yes | Yes | Good |
| Yamazaki H et al, 2022 | Yes | Yes | Yes | Good |
