## Appendix C single group for "Pregnancy and the prognosis of patients previously treated for differentiated thyroid cancer: a systematic review and meta-analysis"

**Appendix C. Quality Assessment –Single Group Studies, Pregnancy**

| **Study, Year** | **Accurate ascertainment confounders** | **Outcomes prespecified and defined, and ascertained using accurate methods?** | **Quality Rating** |
| --- | --- | --- | --- |
| Leboeuf R et al, 2007 | Yes | Unclear | Poor |
| Rosario PW et al, 2007 | Unclear | Unclear | Poor |
| Hirsch D et al, 2010 | Yes | Yes | Fair |
| Rakhlin L et al, 2017 | Yes | Yes | Fair |
| Nobre GM et al, 2021 | Yes | Yes | Fair |
| Colombo C et al, 2022 | Yes | Unclear | Poor |
